# Heterogeneity and district-level factors associated with COVID-19 mortality during three epidemic waves in Indonesia: a nationwide ecological study

**DOI:** 10.1101/2022.06.20.22276672

**Authors:** Henry Surendra, Danarastri Paramita, Nora N Arista, Annisa I Putri, Akbar A Siregar, Evelyn Puspaningrum, Leni Rosylin, Dida Gardera, Montty Girianna, Iqbal RF Elyazar

## Abstract

**Introduction:** Ensuring health equity, especially for vulnerable populations in less developed settings with poor health system is essential for the current and future global health threats. This study examined the heterogeneity of COVID-19 mortality and its association with population health characteristics, health care capacity in responding pandemic, and socio-economic characteristics across 514 districts in Indonesia.

**Methods:** This nationwide ecological study included aggregated COVID-19 cases data from all 514 districts in Indonesia, recorded in the National COVID-19 Task Force database, during the first two years of the epidemic, from 1 March 2020 to 27 February 2022. We calculated incidence and mortality rate by time, sex, and age. We extracted district-level socio-demographics, population health, and health care capacity data from government official sources. We used multivariable linear regression to examine factors associated with higher mortality rate.

**Results:** Of total 5,539,333 reported cases, 148,034 (2·7%) died, and 5,391,299 (97.4%) were recovered. The national mortality rate was 55 per 100,000 population, ranged from 13 per 100,000 population in Papua to 156 per 100,000 population in East Kalimantan province. At district-level, higher mortality rate was associated with higher COVID-19 incidence (p<0.0001), higher proportion of ≥60 years old population (p<0.0001), higher prevalence of diabetes mellitus (p<0.0001), lower prevalence of obesity (p=0.019), lower number of doctors per population (p=0.001), higher life expectancy at birth (p=0.035), and lower formal education (p=0.021). There was no association between COVID-19 mortality rate with expenditure, prevalence of hypertension and pneumonia, vaccine coverage for ≥60 years old population, number of nurses, midwives, and hospitals per population (p>0.05 each).

**Conclusion:** COVID-19 mortality rate in Indonesia was highly heterogeneous and associated with different prevalence of pre-existing comorbidity, healthcare capacity in responding the pandemic, and socio-economic characteristics. This study revealed the need of health capacity strengthening and better resource allocation to ensure optimal health outcomes for vulnerable population.

**What is already known on this topic:**

- The severity of COVID-19 illness and clinical outcomes can be affected by the concentration of comorbidities in susceptible groups in communities, and through disparities of access to health care for preventive measures or prompt diagnosis and treatment.
- However, evidence on the heterogeneity of COVID-19 impact from low- and middle-income country (LMIC) where differences in age distribution, comorbidities, access to quality health services, and other factors, may greatly influence mortality risk, are limited.

**What this study adds:**

- This study affirmed that COVID-19 disproportionately affected areas with high proportion of elder population, high prevalence of diabetes mellitus, lower doctor to population ratio, higher life expectancy at birth, and lower level of formal education.
- These findings indicate that vulnerability to death associated with COVID-19 in LMIC includes not only elder and comorbid, but also males and communities living in area with lower health care capacity and with lower level of education.

**How this study might affect research, practice and/or policy:**

- These findings may inform decisions on health resource allocation against COVID-19 delivering the greatest possible health dividends by prioritising interventions, including even distribution of essential health care need such as doctors, and a tailored risk communication and community engagement for the most vulnerable communities in LMIC, especially with decentralised health systems like in Indonesia.

## Background

Globally, coronavirus disease 2019 (COVID-19) pandemic has caused millions of deaths [1], public health systems collapse [2], and socio-economic disruptions [3,4]. As of 17 June 2022, there have been 535,863,950 total confirmed cases and 6,314,972 deaths reported to the World Health Organization (WHO) [1]. It was reported that employment loss rates have been high since the beginning of the pandemic, and significantly higher among women than men (26% vs 20% in September 2021), with a steadily decreasing trend worldwide [3]. Income loss was reported at 58·4%, with higher rates seen in less developed settings such as Sub-Saharan Africa, Southeast Asia, east Asia, and Oceania [3]. As per 15 June 2022, 11,902,271,619 vaccine doses have been administered [1], yet the pandemic continues to affect the global population, especially the most vulnerable communities.

Ensuring health equity, especially for vulnerable populations in less developed settings with poor health systems is essential for the current COVID-19 pandemic and future global health threats [5-10]. At community-level, it is known that the severity of illness and clinical outcomes can be affected by the concentration of comorbidities in susceptible groups in communities [11-14], and through disparities in access to health care for preventive measures or prompt diagnosis and treatment [14,15]. Recent findings in US, Chile and Brazil suggested that COVID-19 poor outcome was concentrated in groups with higher socio-demographics and health system vulnerability [14, 16-20]. However, the heterogeneity of COVID-19 impacts from low- and middle-income countries (LMIC) where differences in age distribution, comorbidities, access to quality health services, and other factors, may significantly influence mortality risk, are limited.

Indonesia has suffered the highest number of COVID-19 confirmed cases and deaths in Southeast Asia, second only to India in all of Asia [21], at 6,068,075 cases and 156,687 deaths (2·6% case fatality rate (CFR)) up to 19 June 2022 [22]. The first SARS-CoV-2 epidemic wave occurred from 2 March 2020 to 30 April 2021, and a more intense second wave dominated by Delta variant peaked in July 2021 [23], followed by the third wave peaked in February 2022 [22]. The majority of cases and deaths in Indonesia were reported in Java Island, a more developed setting populated by 152 million individuals (56% of the total Indonesia’s population). Recent studies from Indonesia’s capital city of Jakarta suggested that COVID-19 disproportionately affected individuals with older age and pre-existing chronic comorbidities, as well as those areas with lower vaccine coverage, and higher poverty and population density [24,25]. However, data on the impact of COVID-19 across 514 districts of Indonesia that have different pre-existing burdens of major infectious diseases such as malaria, tuberculosis, HIV and other tropical infections [26], as well as non-communicable diseases like cardiovascular diseases, cancers, chronic pulmonary diseases, diabetes, and others [27], are scarce.

Indonesia is the fourth most populous country (population 270 million) and the LMIC featuring great geographic, cultural and socio-economic diversity across the archipelago. The 2020 human development index (HDI) ranged from 0·32 in Nduga District, Papua to 0·87 in Yogyakarta city, Yogyakarta [28]. In addition, substantial proportions of the Indonesian population face barriers in accessing quality health care services due to under-resourced and fragile health systems [26]. A heavily decentralised health systems [30] has resulted in distinct public health capacity across 514 districts of Indonesia. For example, the 2018 Public Health Development Index (PHDI) ranges from 35% in Paniai district, Papua province to 75% in Gianyar district, Bali province [31]. That heterogeneity and the large number of COVID-19 cases and deaths provides insights directly relevant to the national public health response to the COVID-19 crisis, and other LMIC settings. In this study, we assessed the COVID-19 heterogeneity, district-level socio-economics, population health-related conditions, and health care capacity among 514 districts of Indonesia and how those factors were associated with COVID-19 mortality rate during the first 24 months of the epidemic in Indonesia (March 2020 through February 2022).

## Methods

### Study design and participants

This was a nationwide ecological study to assess COVID-19 burden, heterogeneity, and factors associated with mortality rate in all 514 districts in Indonesia. The study analysed aggregated data of individuals diagnosed with COVID-19 based on either rapid antigen diagnostic test (Ag RDT) or polymerase chain reaction (PCR) recorded by the National COVID-19 Task Force from 1 March 2020 to 27 February 2022. In accordance with Indonesia’s national COVID-19 guideline, individuals are categorized as COVID-19 confirmed cases if tested positive by Ag RDT or PCR.

The main outcome variable in this study was district-level COVID-19 mortality rate per 100,000 population. The determinants assessed were gender, proportion of ≥60 years old individuals, prevalence of hypertension, prevalence of diabetes mellitus, prevalence of obesity, prevalence of pneumonia, COVID-19 vaccine coverage for ≥60 years old population, number of doctors, nurses, midwives, and hospitals per 10,000 population, expenditure, life expectancy at birth, and length of formal education.

### Patient and public involvement

This study did not include patients and public in the design, or conduct, or reporting, or dissemination plans. This study was a secondary analysis of aggregated routine surveillance data conducted under a formal collaboration between the National COVID-19 Task Force (Komite Pengendalian COVID-19 dan Pemulihan Ekonomi Nasional) and United Nation Development Program, Indonesia.

### Data collection

The aggregated data of weekly number of COVID-19 cases, incidence rate per 100,000 population, number of COVID-19 deaths among confirmed cases, mortality rate per 100,000 population, by district, sex, and age group from 1 March 2020 to 27 February 2022 were collected from the government official COVID-19 database managed by the National COVID-19 Task Force.

District-level data were collected from various government sources. Data on the number of populations were collected from Statistics Bureau Database available in each province. Data on the number of doctors, nurses, midwives, and hospitals per October 2021 were collected from the Indonesia Ministry of Health records. Data on prevalence of hypertension, diabetes mellitus, obesity, and pneumonia were collected from the National Public Health Development Index 2018 Report [31]. Data on the COVID-19 two-dose vaccine coverage for ≥60 years old population by district per 27 February 2022 were collected from the National COVID-19 Vaccination Database. Data on expenditure, life expectancy at birth, and length of formal education were collected from the Human Development Index 2020 Report [28].

### Statistical analysis

Incidence and mortality rate per 100,000 population was calculated by time (weekly), sex, and province. Male to female incidence and mortality rate ratio (RR) was calculated to compare risk between males and females, and by province. Maps showing spatial distribution of male to female incidence and mortality rate ratio were generated using QGIS 3.20 software.

District-level incidence and mortality rate per 100,000 population, proportion of ≥60 years old population, prevalence of hypertension (%), prevalence of diabetes mellitus (%), prevalence of obesity (%), prevalence of pneumonia (%), COVID-19 vaccine coverage for ≥60 years old population (%), number of doctors, nurses, midwives, and hospitals per 100,000 population, expenditure (millions IDR), life expectancy at birth (years), and length of formal education (years) were calculated then categorised into quartiles. Descriptive statistics included proportions and the chi-squared test to compare district-level characteristics between different quartiles. Spearman’s correlation tests were done to assess correlation between each district-level variable. We used bivariable and multivariable linear regression models to determine factors associated with higher mortality rate at district level, expressed as beta, with 95% confidence intervals. All independent variables with p-value <0.10 in bivariable analysis were included in the multivariable models. Final model selection was informed by likelihood ratio tests. We set statistical significance at 0·05, and all tests were two-sided. All analyses were done in Stata/IC 15.1 (StataCorp, College Station, TX, USA). This study is reported as per Strengthening the Reporting of Observational Studies in Epidemiology (STROBE) guidelines [31].

### Ethics

This study was a secondary analysis of aggregated routine surveillance data collected by COVID-19 control program, with district as the analysis unit. Use of the data, which was aggregated, and at the population level is permitted by the Indonesian Ministry of Health under Regulation Number 45 (2014), Article 3, paragraphs 1 and 2. There was no ethical approval sought for this study.

## Results

### Cumulative COVID-19 incidence and mortality rate

Table 1 presents the reported cumulative COVID-19 incidence and mortality rate in Indonesia between 1 March 2020 and 27 February 2022. A total of 5,539,333 COVID-19 cases were recorded on the Indonesia National COVID-19 Database. Of those, 148,034 (2·7%) were deceased, and 5,391,299 (97.4%) were recovered. The overall incidence and mortality rates were 2,050 and 55 per 100,000 population, respectively. The incidence and mortality rate varied widely by province. The highest incidence rate was reported in DKI Jakarta (10,764 cases per 100,000 population), and the lowest was reported in West Nusa Tenggara (667 cases per 100,000 population). The highest mortality rate was reported in East Kalimantan (156 per 100,000 population), while the lowest was reported in Papua (13 per 100,000 population).

**Table 1.** Cumulative COVID-19 incidence and mortality rate between March 2020 and February 2022 in Indonesia, stratified by province.

| Region/Province | Total cases | Incidence per 100,000 population | Total deaths | Mortality per 100,000 population |
| --- | --- | --- | --- | --- |
| **Overall** | **5,539,333** | **2,050.06** | **148,034** | **54.79** |
| **Sumatera** |  |  |  |  |
| Aceh | 40,302 | 768.06 | 2,087 | 39.77 |
| North Sumatera | 139,683 | 939.05 | 2,954 | 19.86 |
| West Sumatera | 98,913 | 1,792.15 | 2,191 | 39.77 |
| Riau | 141,166 | 2,324.06 | 4,170 | 68.65 |
| Jambi | 33,924 | 971.10 | 798 | 22.84 |
| South Sumatera | 76,343 | 929.02 | 3,162 | 38.48 |
| Bengkulu | 26,951 | 1,347.86 | 482 | 24.11 |
| Lampung | 64,718 | 711.53 | 3,952 | 43.45 |
| Bangka Belitung Islands | 58,493 | 4,293.34 | 1,487 | 107.77 |
| Riau Islands | 63,683 | 3,300.66 | 1,772 | 91.84 |
| **Java** |  |  |  |  |
| DKI Jakarta | 1,167,506 | 10,764.25 | 14,585 | 134.47 |
| West Java | 999,997 | 2,214.28 | 15,048 | 33.32 |
| Central Java | 573,628 | 1,577.46 | 30,976 | 85.18 |
| Yogyakarta | 190,336 | 5,241.95 | 5,370 | 147.89 |
| East Java | 531,169 | 1,312.21 | 30,296 | 74.84 |
| Banten | 265,041 | 2,471.85 | 2,829 | 26.38 |
| **Bali and Nusa Tenggara** |  |  |  |  |
| Bali | 152,466 | 3,616.22 | 4,390 | 104.12 |
| West Nusa Tenggara | 35,133 | 666.63 | 863 | 16.37 |
| East Nusa Tenggara | 73,736 | 1,362.62 | 1,365 | 25.22 |
| **Kalimantan** |  |  |  |  |
| West Kalimantan | 52,597 | 969.92 | 1,078 | 19.88 |
| Central Kalimantan | 52,114 | 2,027.55 | 1,454 | 56.57 |
| South Kalimantan | 81,842 | 2,034.33 | 2,462 | 61.20 |
| East Kalimantan | 187,834 | 5,287.84 | 5,525 | 155.54 |
| North Kalimantan | 38,752 | 5,976.49 | 813 | 125.38 |
| **Sulawesi** |  |  |  |  |
| North Sulawesi | 48,412 | 1,832.48 | 1,103 | 41.75 |
| Central Sulawesi | 52,470 | 1,775.29 | 1,621 | 54.85 |
| South Sulawesi | 134,225 | 1,423.85 | 2,327 | 24.68 |
| Southeast Sulawesi | 24,529 | 930.73 | 545 | 20.68 |
| Gorontalo | 12,854 | 1,088.72 | 462 | 39.13 |
| West Sulawesi | 13,723 | 879.69 | 357 | 22.88 |
| **Maluku and Papua** |  |  |  |  |
| Maluku | 18,382 | 995.18 | 279 | 15.10 |
| North Maluku | 13,490 | 1,031.50 | 308 | 23.55 |
| West Papua | 29,129 | 2,553.61 | 362 | 31.73 |
| Papua | 45,792 | 1,055.03 | 561 | 12.93 |

### Heterogeneity of COVID-19 incidence and mortality rate at province-level

Figure 1 shows the weekly incidence and mortality at the national and province-level. The data suggest that Indonesia had experienced three pandemic waves with the highest mortality recorded in week 73^rd^ (12 to 18 July 2021) where cases were predominantly infected by Delta variant (Figure 1). The province-level data show that DKI Jakarta had consistently experienced the highest incidence rate over time (Figure 2A). Overall, the mortality rate increased over time, with the highest rate seen in East Kalimantan, followed by Yogyakarta, DKI Jakarta, and North Kalimantan (Figure 2B).

**Figure 1.**
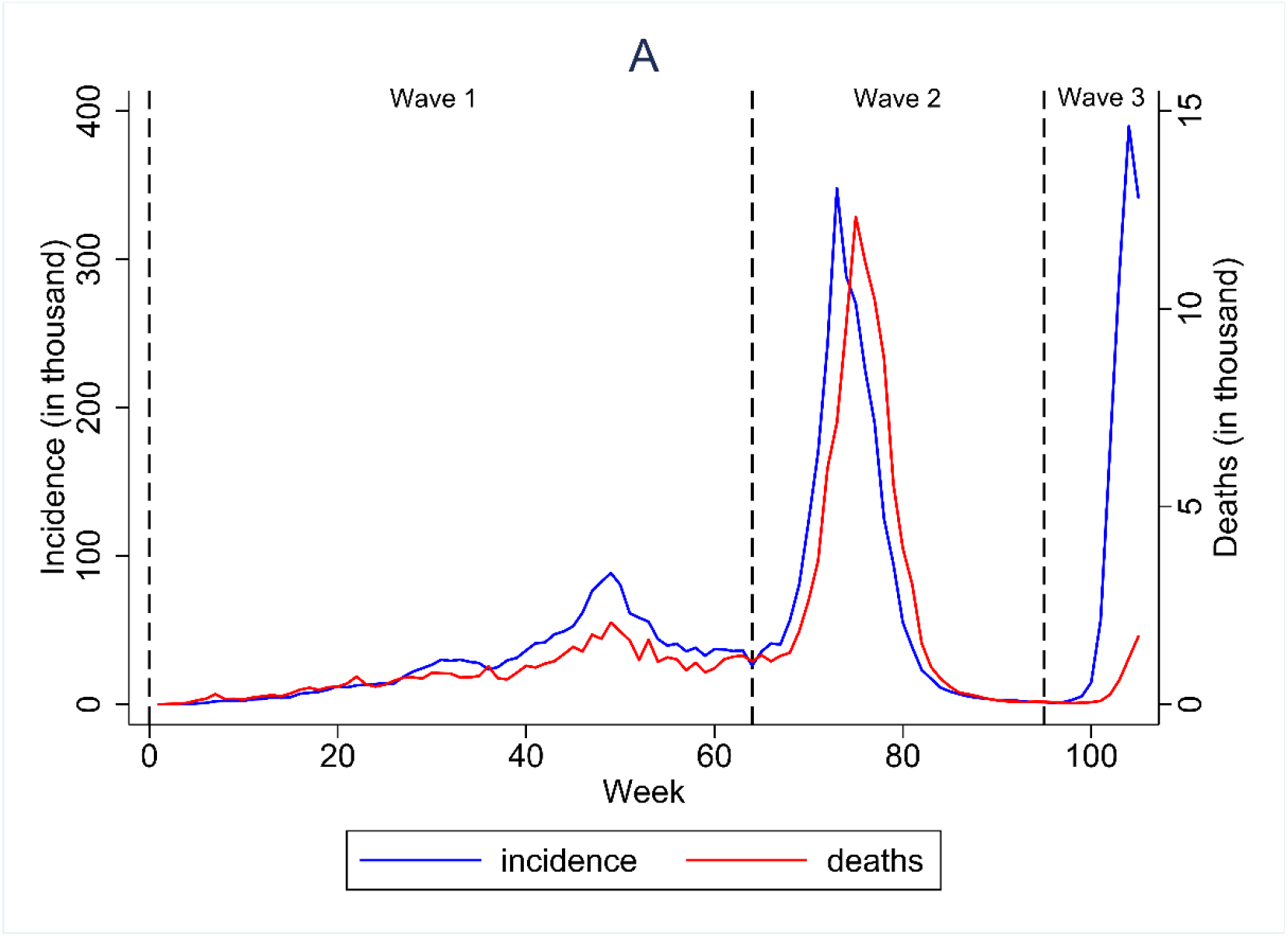
COVID-19 incidence and mortality over the first three epidemic waves in Indonesia between March 2020 to February 2022.

**Figure 2:**
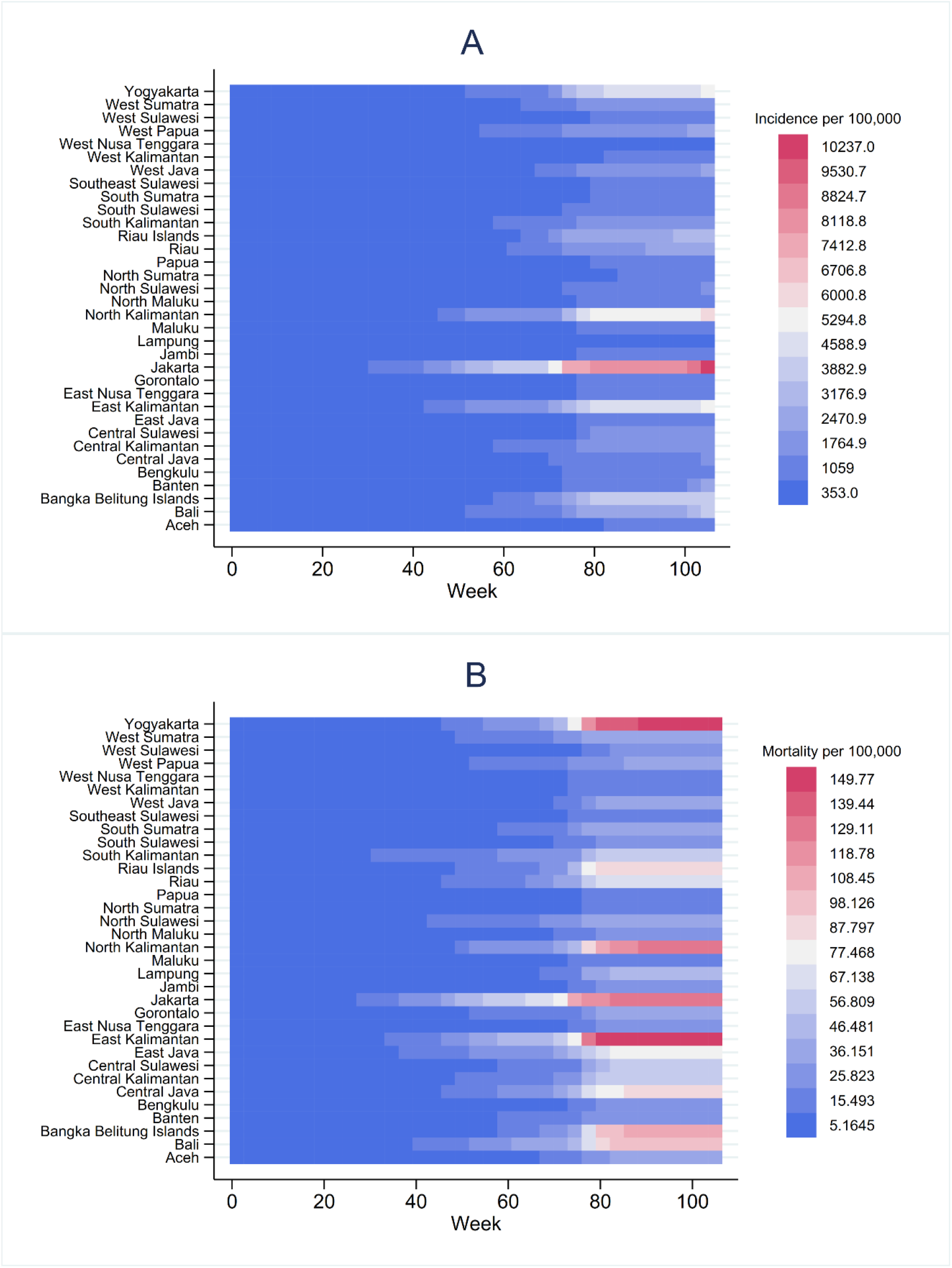
Heatmaps of weekly incidence rate per 100,000 population (A) and mortality rate per 100,000 population by province (B).

The mortality rate was higher for males than females across 34 provinces in Indonesia (Table 2), with the highest male to female mortality rate ratio (RR) seen in East Nusa Tenggara (RR 3.00), followed by Yogyakarta (RR 2.61), Southeast Sulawesi (RR 2.52), Banten (RR 2.51), and West Papua (RR 2.50). The lowest male to female mortality was reported in Aceh province (RR 1.15). The incidence rate was also higher for males than females in the majority of provinces (94.12%), except for in Aceh and Bali (RR 0.92 and 0.99, respectively) (Supplementary Table 1). The highest male to female mortality rate ratio was seen in East Kalimantan (RR 2.42), followed by North Kalimantan (RR 2.17), Riau islands, Papua, and West Papua (RR 2.12 each).

**Table 2.** Overall COVID-19 mortality rate between March 2020 and February 2022 in Indonesia by sex and province.

| Province | Male |  |  | Number of Deaths | Female |  | Male-female mortality rate ratio |
| --- | --- | --- | --- | --- | --- | --- | --- |
|  | Number of Deaths | Number of Population | Mortality per 100,000 population |  | Number of Population | Mortality per 100,000 population |  |
| Aceh | 1114 | 2,691,800 | 41.38 | 973 | 2,696,300 | 36.09 | 1.15 |
| Bali | 2664 | 2,221,400 | 119.92 | 1719 | 2,193,100 | 78.38 | 1.53 |
| Banten | 1570 | 6,557,900 | 23.94 | 1232 | 12,895,300 | 9.55 | 2.51 |
| Bengkulu | 236 | 1,015,200 | 23.25 | 237 | 1,994,300 | 11.88 | 1.96 |
| Yogyakarta | 3020 | 1,935,400 | 156.04 | 2346 | 3,919,200 | 59.86 | 2.61 |
| Jakarta | 7734 | 5,267,800 | 146.81 | 6727 | 10,576,400 | 63.60 | 2.31 |
| Gorontalo | 245 | 593,500 | 41.28 | 217 | 1,186,300 | 18.29 | 2.26 |
| Jambi | 401 | 1,831,300 | 21.90 | 377 | 3,604,200 | 10.46 | 2.09 |
| West Java | 7666 | 25,111,200 | 30.53 | 7357 | 49,565,200 | 14.84 | 2.06 |
| Central Java | 15,750 | 17,237,300 | 91.37 | 14,192 | 34,738,200 | 40.85 | 2.24 |
| East Java | 14,962 | 19,722,200 | 75.86 | 14,907 | 39,955,900 | 37.31 | 2.03 |
| West Kalimantan | 593 | 2,591,400 | 22.88 | 481 | 5,104,900 | 9.42 | 2.43 |
| South Kalimantan | 1355 | 2,156,700 | 62.83 | 1096 | 4,268,600 | 25.68 | 2.45 |
| Central Kalimantan | 791 | 1,394,900 | 56.71 | 659 | 2,686,300 | 24.53 | 2.31 |
| East Kalimantan | 2946 | 1,902,900 | 154.82 | 2566 | 3,664,700 | 70.02 | 2.21 |
| North Kalimantan | 405 | 374,100 | 108.26 | 402 | 708,400 | 56.75 | 1.91 |
| Bangka Belitung Islands | 706 | 756,900 | 93.28 | 780 | 1,469,800 | 53.07 | 1.76 |
| Riau Islands | 963 | 1,179,000 | 81.68 | 809 | 2,309,500 | 35.03 | 2.33 |
| Lampung | 1887 | 4,364,300 | 43.24 | 2061 | 8,534,800 | 24.15 | 1.79 |
| Maluku | 146 | 900,400 | 16.22 | 131 | 1,787,100 | 7.33 | 2.21 |
| North Maluku | 137 | 637,300 | 21.50 | 151 | 1,252,300 | 12.06 | 1.78 |
| West Nusa Tenggara | 445 | 2,563,900 | 17.36 | 416 | 5,225,900 | 7.96 | 2.18 |
| East Nusa Tenggara | 811 | 2,731,600 | 29.69 | 545 | 5,513,400 | 9.89 | 3.00 |
| Papua | 291 | 1,777,700 | 16.37 | 229 | 3,393,100 | 6.75 | 2.43 |
| West Papua | 205 | 518,100 | 39.57 | 156 | 986,000 | 15.82 | 2.50 |
| Riau | 2154 | 3,553,200 | 60.62 | 2016 | 6,951,200 | 29.00 | 2.09 |
| West Sulawesi | 182 | 692,200 | 26.29 | 175 | 1,378,100 | 12.70 | 2.07 |
| South Sulawesi | 1251 | 4,348,500 | 28.77 | 1066 | 8,888,800 | 11.99 | 2.40 |
| Central Sulawesi | 810 | 1,565,100 | 51.75 | 804 | 3,081,700 | 26.09 | 1.98 |
| Southeast Sulawesi | 304 | 1,352,900 | 22.47 | 241 | 2,703,500 | 8.91 | 2.52 |
| North Sulawesi | 601 | 1,279,600 | 46.97 | 499 | 2,512,900 | 19.86 | 2.37 |
| West Sumatera | 1112 | 2,760,600 | 40.28 | 1055 | 5,545,700 | 19.02 | 2.12 |
| South Sumatera | 1610 | 4,358,000 | 36.94 | 1543 | 8,600,800 | 17.94 | 2.06 |
| North Sumatera | 1578 | 7,392,700 | 21.35 | 1364 | 14,798,400 | 9.22 | 2.32 |

Number of populations by sex was obtained from the projected number of populations in 2020 which are publicly available from https://www.bps.go.id/indicator/12/1886/1/jumlah-penduduk-hasil-proyeksi-menurut-provinsi-dan-jenis-kelamin.html

### Heterogeneity and factors associated with higher mortality rate at district-level

Table 3 presents the summary of district-level COVID-19 burden, prevalence of health-related conditions, and health care capacity associated with COVID-19 mortality rate across 514 districts in Indonesia. The incidence and mortality rate were highly heterogeneous, ranged from 8.13 to 10,626.09 per 100,000 population, and 0.28 to 283.82 per 100,000 population, respectively. Compared to eastern Indonesia, the mortality rate tends to be higher in the western areas, especially in districts and cities located in Java and Kalimantan Island (Figure 3A). Higher incidence rate was seen in Kalimantan, Sulawesi, and Papua Island (Figure 3B). Based on quartiles, the mortality rates ranged from 0.28 to 16.43 per 100,000 population (quartile 1), 16.43 to 31.47 per 100,000 population (quartile 2), 31.47 to 64.68 per 100,000 population (quartile 3), and 64.68 to 283.82 per 100,000 population (quartile 4). Of 129 districts in the highest mortality rate quartile, 71.32% had the highest incidence rate.

**Table 3.**
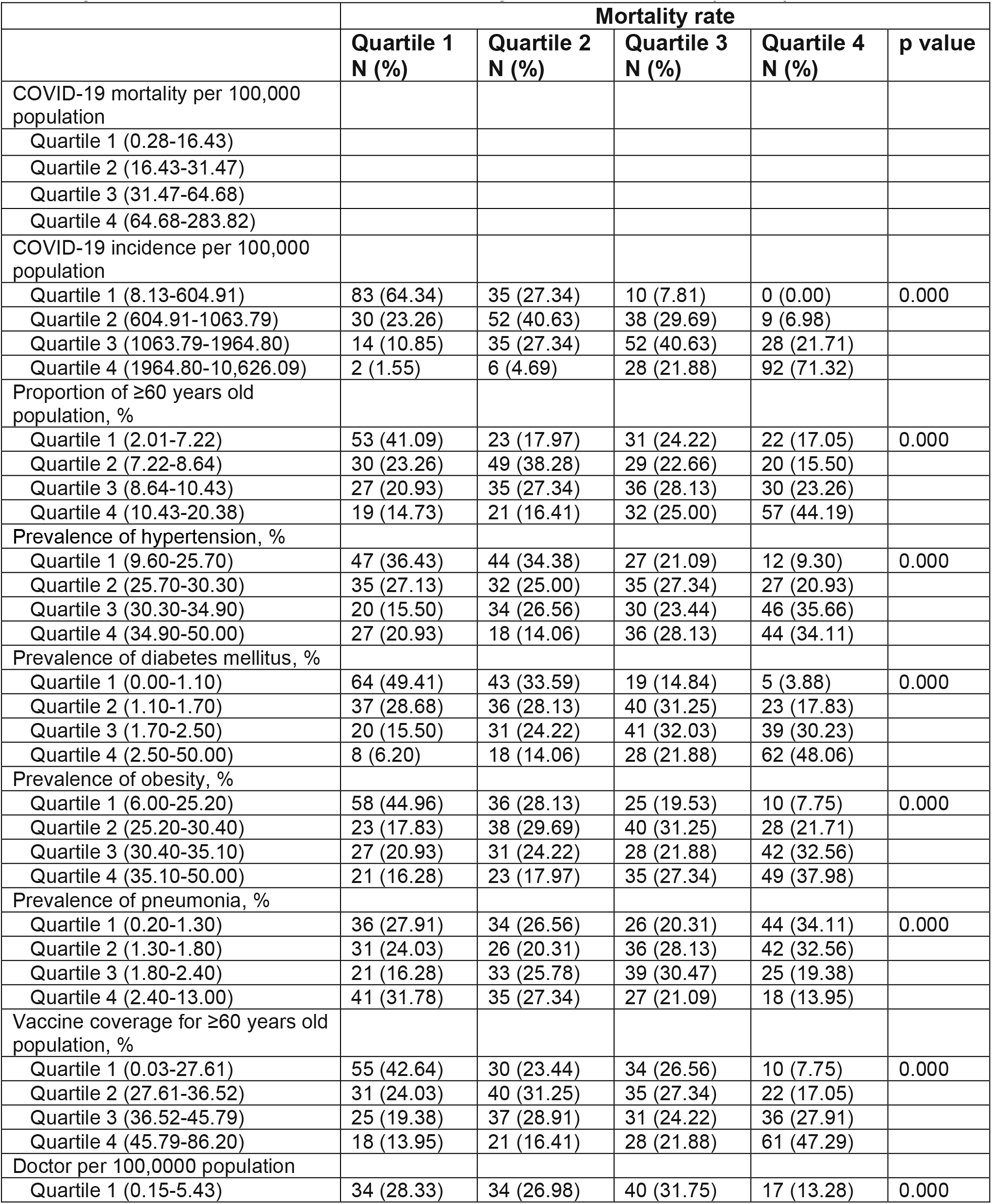
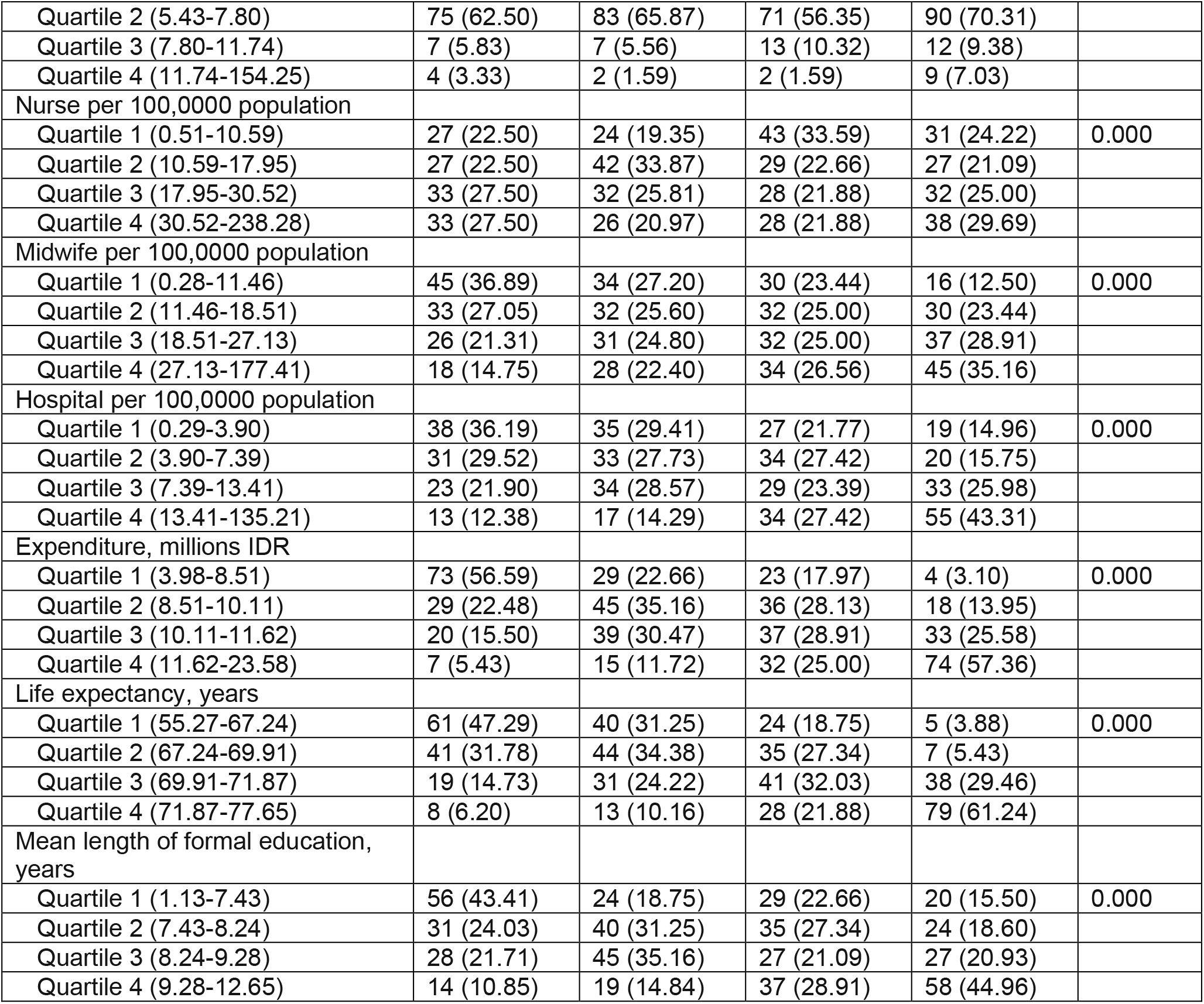
Summary of COVID-19 burden, prevalence of health-related conditions, health care capacity, and socio-economic characteristics associated with higher COVID-19 mortality rate between March 2020 and February 2022 in Indonesia (N=514)

**Figure 3:**
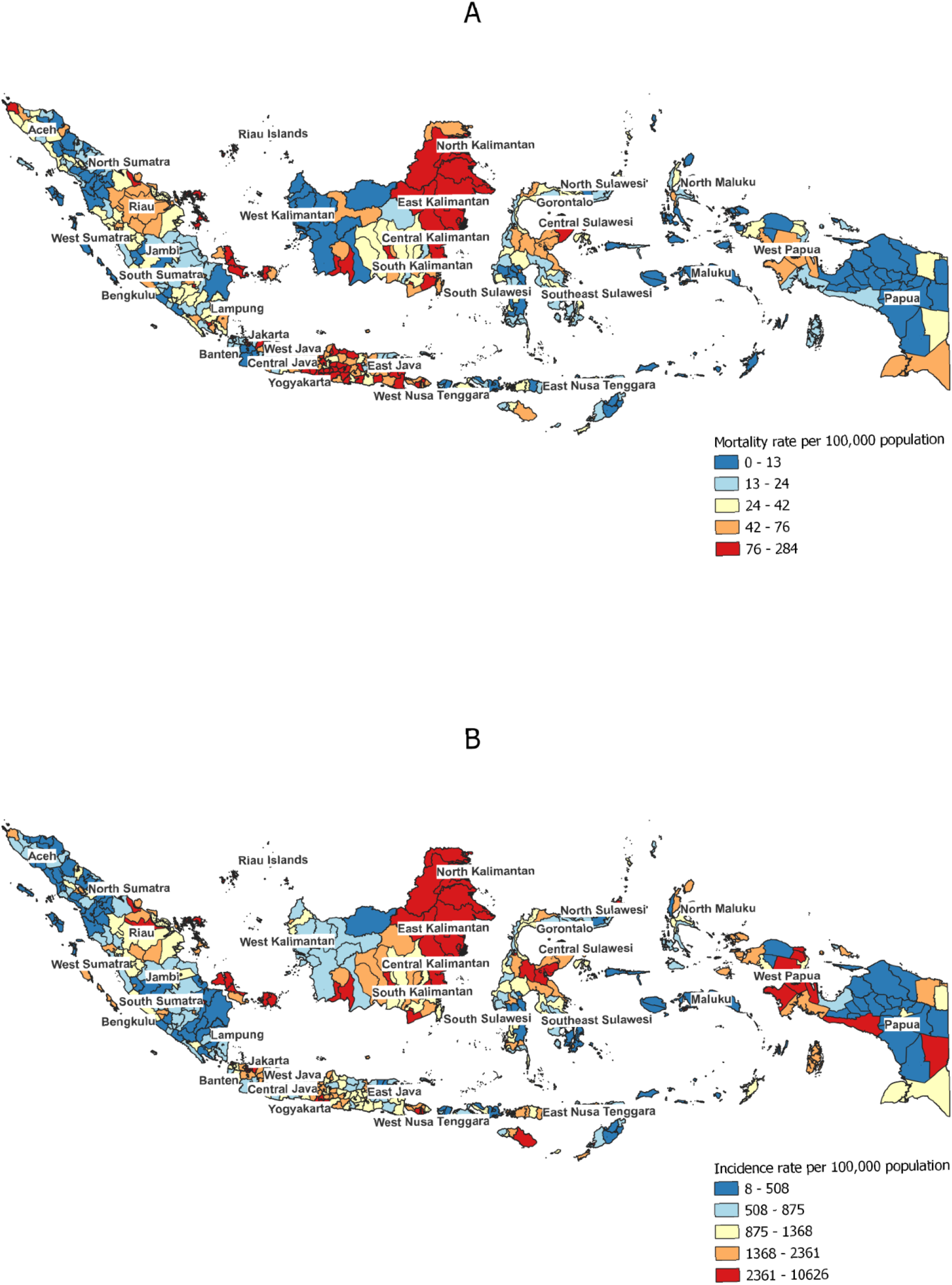
District-level mortality rate (A), and incidence rate (B) in Indonesia between March 2020 and February 2022.

Regarding population health-related conditions, 44.19% of districts with the highest mortality rate had the highest proportion of ≥60 years old population (10.43-20.38%), 34.11% had the highest prevalence of hypertension (34.90-50.00%), 48.06% had the highest prevalence of diabetes mellitus (2.50-50.00%), 37.98% had the highest prevalence of obesity (35.10-50.00%), and 13.95% had the highest prevalence of pneumonia (2.40-13.00%). Regarding health care capacity, 47.29% of districts with the highest mortality rate had the highest COVID-19 vaccine coverage for ≥60 years old population (45.79-86.20%), 13.28% had the lowest number of doctors per 100,000 population (0.15-5.43), 24.22% had the lowest number of nurses per 100,000 population (0.51-10.59), 12.50% had the lowest number of midwives per 100,000 population (0.28-11.46), and 43.31% had the lowest number of hospitals per 100,000 population (0.29-3.90). Regarding socio-economic, only 3.10% of districts with the highest mortality rate had the lowest expenditure (IDR 3.98-8.51 million), 3.88% had the lowest life expectancy at birth (55.27-67.24 years), and 15.50% had the lowest mean of length of formal education (1.13-7.43 years) (See Table 3 for details).

In bivariable linear regression analysis (Table 4), district-level COVID-19 mortality rate was associated with COVID-19 incidence rate, proportion of ≥60 years old individuals, prevalence of hypertension, prevalence of diabetes mellitus, prevalence of obesity, prevalence of pneumonia, vaccine coverage for ≥60 years old population, doctors per population, hospital per population, expenditure, life expectancy, and length of formal education. We found no association between mortality rate with number of nurses and midwives per population. Correlation matrix of COVID-19 burden, prevalence of health-related conditions, vaccine coverage for >60 years old population, health care capacity, and socio-economic characteristics can be seen in Figure 4.

**Table 4.**
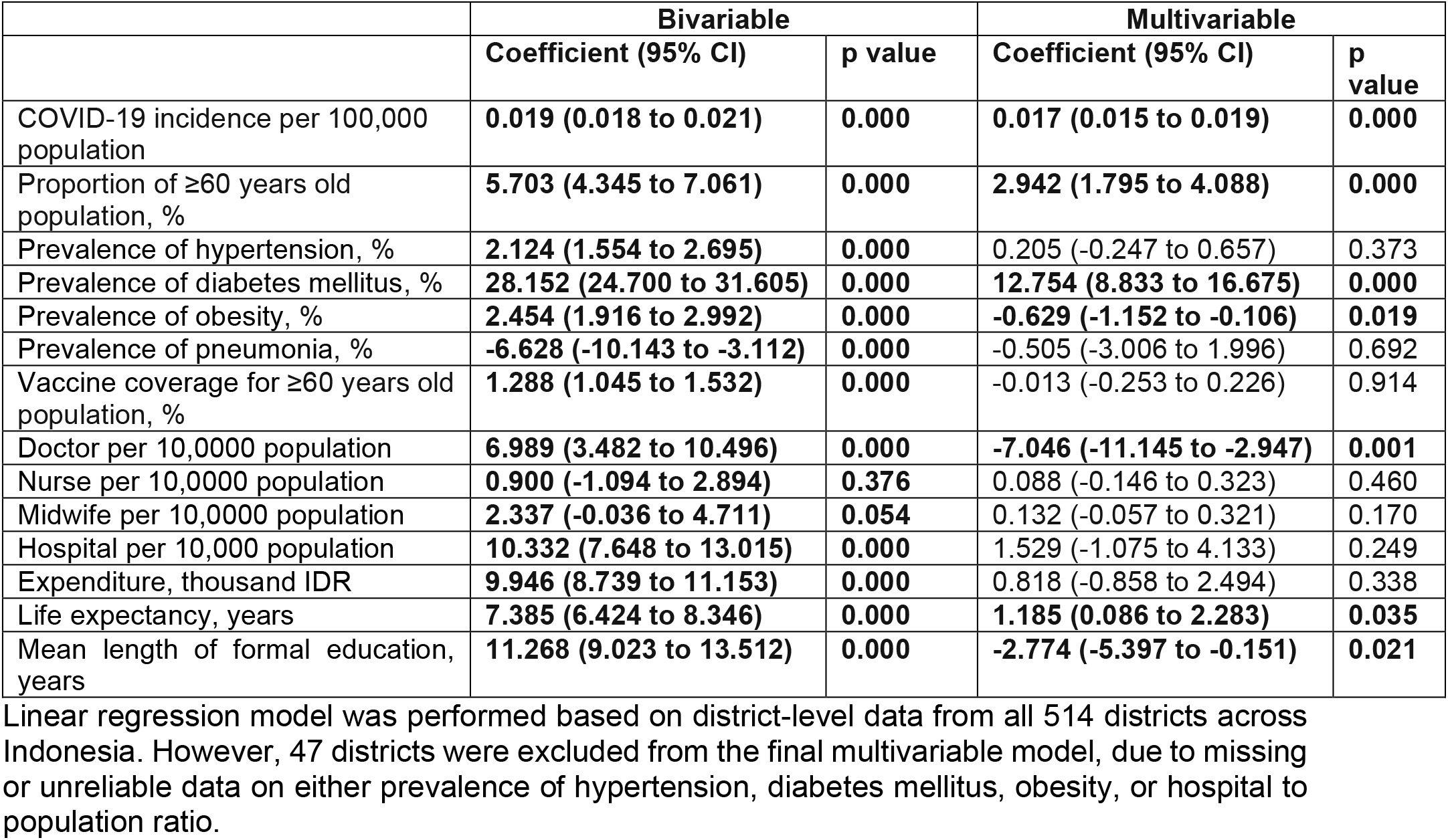
District-level risk factors associated with higher COVID-19 mortality rate between March 2020 and February 2022 in Indonesia.

|  | Bivariable |  | Multivariable |  |
| --- | --- | --- | --- | --- |
|  | Coefficient (95% CI) | p value | Coefficient (95% CI) | p value |
| COVID-19 incidence per 100,000 population | **0.019 (0.018 to 0.021)** | **0.000** | **0.017 (0.015 to 0.019)** | **0.000** |
| Proportion of $\geq 60$ years old population, % | **5.703 (4.345 to 7.061)** | **0.000** | **2.942 (1.795 to 4.088)** | **0.000** |
| Prevalence of hypertension, % | **2.124 (1.554 to 2.695)** | **0.000** | 0.205 (-0.247 to 0.657) | 0.373 |
| Prevalence of diabetes mellitus, % | **28.152 (24.700 to 31.605)** | **0.000** | **12.754 (8.833 to 16.675)** | **0.000** |
| Prevalence of obesity, % | **2.454 (1.916 to 2.992)** | **0.000** | **-0.629 (-1.152 to -0.106)** | **0.019** |
| Prevalence of pneumonia, % | **-6.628 (-10.143 to -3.112)** | **0.000** | -0.505 (-3.006 to 1.996) | 0.692 |
| Vaccine coverage for $\geq 60$ years old population, % | **1.288 (1.045 to 1.532)** | **0.000** | -0.013 (-0.253 to 0.226) | 0.914 |
| Doctor per 10,000 population | **6.989 (3.482 to 10.496)** | **0.000** | **-7.046 (-11.145 to -2.947)** | **0.001** |
| Nurse per 10,000 population | **0.900 (-1.094 to 2.894)** | **0.376** | 0.088 (-0.146 to 0.323) | 0.460 |
| Midwife per 10,000 population | **2.337 (-0.036 to 4.711)** | **0.054** | 0.132 (-0.057 to 0.321) | 0.170 |
| Hospital per 10,000 population | **10.332 (7.648 to 13.015)** | **0.000** | 1.529 (-1.075 to 4.133) | 0.249 |
| Expenditure, thousand IDR | **9.946 (8.739 to 11.153)** | **0.000** | 0.818 (-0.858 to 2.494) | 0.338 |
| Life expectancy, years | **7.385 (6.424 to 8.346)** | **0.000** | **1.185 (0.086 to 2.283)** | **0.035** |
| Mean length of formal education, years | **11.268 (9.023 to 13.512)** | **0.000** | **-2.774 (-5.397 to -0.151)** | **0.021** |

Linear regression model was performed based on district-level data from all 514 districts across Indonesia. However, 47 districts were excluded from the final multivariable model, due to missing or unreliable data on either prevalence of hypertension, diabetes mellitus, obesity, or hospital to population ratio.

**Figure 4:**
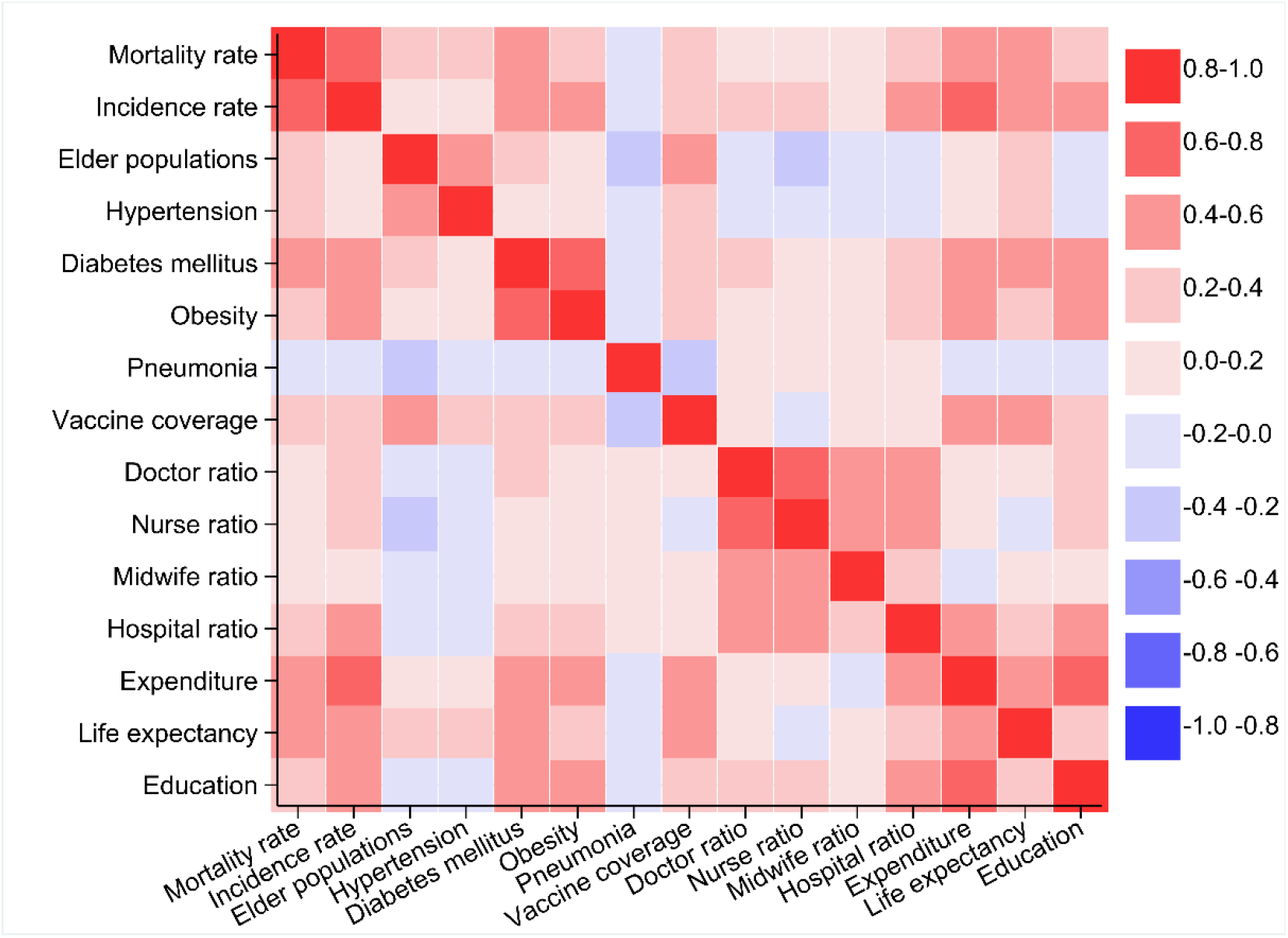
Correlation matrix of COVID-19 burden, prevalence of health-related conditions, vaccine coverage for >60 years old population, health care capacity, and socio-economic characteristics. Significance level of the correlation can be seen in Supplementary Table 2.

In the multivariable model, higher mortality rate was associated with higher COVID-19 incidence (p<0.0001), higher proportion of ≥60 years old population (p<0.0001), higher prevalence of diabetes mellitus (p<0.0001), lower prevalence of obesity (p=0.019), lower number of doctors per population (p=0.001), higher life expectancy at birth (p=0.035), and shorter length of formal education (p=0.021). There was no association between mortality rate with prevalence of hypertension, prevalence of pneumonia, vaccine coverage for ≥60 years ole population, number of nurses, midwives, and hospitals per population, and expenditure (p>0.05 each).

## Discussion

This nationwide ecological study described the epidemiological surveillance data of confirmed COVID-19 cases reported from 514 districts, during three epidemic waves in the first 24 months of the SARS-CoV-2 transmission in Indonesia. The overall case fatality rate was 2·7% (148,034/5,539,333), which equate to a mortality rate of 55 per 100,000 population. The mortality rate was highly heterogeneous and increased over time, with the highest rate seen in East Kalimantan, followed by Yogyakarta, DKI Jakarta, and North Kalimantan. The mortality rate tends to be higher for males than female in the majority of provinces in Indonesia, with the highest male-to-female mortality rate ratio seen in eastern Indonesia provinces. At district-level, higher mortality rate was associated with higher COVID-19 incidence, higher proportion of ≥60 years old population, higher prevalence of diabetes mellitus, lower prevalence of obesity, lower number of doctors per population, higher life expectancy at birth, and shorter length of formal education.

Findings from this study revealed the extent of health inequity and its association with COVID-19 mortality across 514 districts in Indonesia. In contrast to a previous province-level analysis (n=34) found the absence of correlation between mortality rate with doctor to population ratio during the first six months of the epidemic [33], our district-level analysis found that a higher COVID-19 mortality rate was strongly associated with a lower number of doctors per 100,000 population at district-level. This finding contrasts a previous finding from a higher-income country, reporting an association between higher COVID-19 mortality rate with higher physicians density in France [34]. In Indonesia, the lack of doctors, combined with the high number of infections [35] and deaths among health care workers and doctors [36], especially during the early phase of the epidemic, had weakened the health system resilience. In the context of a heavily decentralized health system such as in Indonesia [30], coordination and prioritisation of available resources and public health intervention will be critical to ensure optimal health outcomes for vulnerable communities, especially for those areas with lower number of doctors per population. Short-term solutions such as reallocating doctors to districts with the lowest doctor to population ratio may assist local government in mitigating this inequity in health care access. Improving access to medical doctor training for local residents, and providing incentives to doctors in rural areas can provide long-term solution in ensuring future health equity.

Consistent with individual-level risk factors at from diverse settings [18, 25, 37-38], our district-level analysis affirms that higher proportion of elder population (also reflected by higher life expectancy at birth), males gender, and higher prevalence of diabetes mellitus were significantly associated with higher COVID-19 mortality rate in the general population of Indonesia. This was also consistent with findings from a previous study suggesting that individual-level risk factors such as older age, male sex, and pre-existing hypertension and diabetes were associated with an elevated risk of COVID-19 death during the early epidemic phase in DKI Jakarta, Indonesia [25]. These findings indicate the need of controlling both SARS-Cov-2 transmission and non-communicable diseases, especially in areas with high prevalence of diabetes mellitus. Reducing COVID-19 mortality in such settings may necessitate comprehensive and specific interventions such as improving diagnosis and case management of those known non-communicable diseases, community awareness, as well as a sustainable and accessible social security network that may reduce vulnerability of these communities.

Our analysis reveals the heterogeneity of COVID-19 burden across 34 provinces and 514 districts in Indonesia. The province-level findings suggesting that mortality and incidence rate was higher for males than females, may reflect the cultural phenomenon in Indonesia where males were more likely to get more exposure to the SARS-Cov-2 due to their domestic responsibility to work even during the social restrictions were implemented. Our following findings suggesting higher mortality rate among districts with lower-level of formal education (reflected by shorter mean length of formal education) was consistent with findings from more developed settings such as in the US [11, 39] and Sweden [40]. A lower educational level might be associated with a lower health literacy that can lead to lower access and understanding of public health information. A tailored risk communication and community engagement strategy targeting population living in districts with lower education level is warranted in addressing health inequity in the future.

A previous cohort study in DKI Jakarta reported that higher COVID-9-related mortality risk was significantly associated with lower COVID-19 vaccine coverage at subdistrict-level [24]. By contrast, our current study found that there was no association between district-level COVID-9 mortality rate with COVID-19 vaccine coverage among >60 years old population in Indonesia. This absence of association could possibly be explained by the very low vaccine coverage across the 514 districts. The vaccine coverage was highly varied from 0.03% to 86.20%. The majority of districts (82.3%) still had a vaccine coverage of less than 50% after one year of vaccine roll-out (27 February 2022), thus further highlights the extent of inequity across the country. A previous study from Brazil reported that rapid scaling up of vaccination coverage among elderly Brazilians was associated with significant declines in relative mortality compared with younger individuals, in a setting where the gamma variant predominated [41]. Those findings from DKI Jakarta and Brazil, highlight that rapid vaccination roll outs targeting the most vulnerable is crucial in reducing COVID-19-related deaths. As per 19 June 2022, the coverage for two-dose and three-doses COVID-19 vaccination in Indonesia was 80.14% (166,911,457/208,265,720 targeted population) and 21.26% (44,273,456/208,265,720), respectively [42].

This study had several limitations. Firstly, this study was based on routine surveillance data derived from district-level health office case reports. The imperfect contact tracing, testing, and reporting activities could result in underreporting of cases, especially those asymptomatic and mild cases, which could cause an overestimation of case fatality rate and an underestimation of mortality rate per population in this study. Secondly, as this study was based on aggregated data available at district level, our analysis was unable to capture and adjust the effect of important individual-level risk factors. Therefore, interpretation of the study findings should be restricted to district-level risk factors. Finally, previous studies in Indonesia have suggested the association between COVID-19 mortality rate with population density [24] and ratio of hospitals bed [33]. However, these variables were not evaluated in our study, due to limited access to the relevant data.

In conclusion, our analysis suggested that COVID-19 disproportionately affected districts with high proportion of elder population, high prevalence of diabetes mellitus, lower doctor to population ratio, higher life expectancy at birth, and lower level of formal education. These findings indicate that vulnerability to death associated with COVID-19 includes not only elders and comorbid, but also males and communities living in area with lower health care capacity and with lower level of education. These findings may inform decisions on health resource allocation against COVID-19 delivering the greatest possible health dividends by prioritising interventions, including even distribution of essential health care need such as doctors, and a tailored risk communication and community engagement for the most vulnerable communities in LMIC, especially with decentralised health systems like in Indonesia. Future nationwide studies incorporating individual and district-level data to assess vulnerability associated with COVID-19-related morbidity and mortality are needed to better comprehend the COVID-19 impact and to better prioritise interventions for the most vulnerable communities.

## Data Availability

After publication, the datasets used for this study will be made available to others on reasonable requests to the corresponding author, including a detailed research proposal, study objectives and statistical analysis plan.

## Acknowledgment

We acknowledge the Ministry of Health of Republic of Indonesia, the National COVID-19 Task Force (Komite Pengendalian COVID-19 dan Pemulihan Ekonomi Nasional), and all health care workers involved in the care for the COVID-19 patients, as well as those involved in the field data collection.

## Contributors

HS was the principal investigator of this study. HS designed the study, did the analysis, and had full access to all of the data in the study and take responsibility for the integrity of the data and the accuracy of the data analysis. HS, DP, NNA, AIP, AAS, and EP contributed to data collection and verification. LR, DG, and MG supervised the data collection and verification. HS and IRFE drafted the paper. All authors critically revised the manuscript for important intellectual content and all authors gave final approval for the version to be published.

## Acknowledgments

We acknowledge all health care workers involved in the care for the COVID-19 patients, as well as those involved in the field data collection.

## References

[1] World Health Organization. WHO Coronavirus Disease (COVID-19) Dashboard. Available: https://covid19.who.int [Accessed 19 June 2022].

[2] Haldane V, Foo CD, Abdalla SM, et al. Health systems resilience in managing the COVID-19 pandemic: lessons from 28 countries. Nature Med 2021; 27:964–980.

[3] Flor LS, Friedman J, Spencer CN, et al. Quantifying the effects of the COVID-19 pandemic on gender equality on health, social, and economic indicators: a comprehensive review of data from March, 2020, to September, 2021. Lancet 2022.

[4] Barlow P, Schalkwyk MCIv, McKee M, et al. COVID-19 and the collapse of global trade: building an effective public health response. Lancet Planet Heal 2021; 5:e102–07.

[5] Worl Health Organization. Strategic preparedness, readiness and response plan to end the global COVID-19 emergency in 2022. Geneva: World Health Organization; 2022.

[6] Lowcock EC, Rosella LC, Foisy J, et al. The social determinants of health and pandemic h1n1 2009 influenza severity. Am J Public Health 2012;102:51–8.

[7] Mayoral JM, Alonso J, Garín O, et al. Social factors related to the clinical severity of influenza cases in Spain during the A (H1N1) 2009 virus pandemic. BMC Public Health 2013;118:1–7.

[8] Grantz KH, Rane MS, Salje H, et al. Disparities in influenza mortality and transmission related to sociodemographic factors within Chicago in the pandemic of 1918. Proc Natl Acad Sci U S A 2016;113:13839–44.

[9] Mamelund SE. A socially neutral disease? Individual social class, household wealth and mortality from Spanish influenza in two socially contrasting parishes in Kristiania 1918-19. Soc Sci Med 2006;62:923–40.

[10] Fallah MP, Skrip LA, Gertler S, Yamin D, Galvani AP. Quantifying Poverty as a Driver of Ebola Transmission. PLoS Negl Trop Dis 2015;9:1–9.

[11] Seligman B, Ferranna M, Bloom DE. Social determinants of mortality from COVID-19: A simulation study using NHANES. PLoS Med 2021;18:1–13.

[12] Acharya R, Porwal A. A vulnerability index for the management of and response to the COVID-19 epidemic in India: an ecological study. Lancet Glob Heal 2020;8:e1142–51.

[13] Yoshikawa Y, Kawachi I. Association of Socioeconomic Characteristics with Disparities in COVID-19 Outcomes in Japan. JAMA Netw Open 2021;4:1–13.

[14] Rocha R, Atun R, Massuda A, Rache B, Spinola P, Nunes L, et al. Effect of socioeconomic inequalities and vulnerabilities on health-system preparedness and response to COVID-19 in Brazil: a comprehensive analysis. Lancet Glob Heal 2021;9:e782–92.

[15] Harlem G. Descriptive analysis of social determinant factors in urban communities affected by COVID-19. J Public Heal (United Kingdom) 2020;42:466–9.

[16] Karmakar M, Lantz PM, Tipirneni R. Association of Social and Demographic Factors with COVID-19 Incidence and Death Rates in the US. JAMA Netw Open 2021;4:1–12.

[17] Mena GE, Martinez PP, Mahmud AS, Marquet PA, Buckee CO, Santillana M. Socioeconomic status determines COVID-19 incidence and related mortality in Santiago, Chile. Science (80-) 2021;372.

[18] Oliveira EA, Colosimo EA, Simões e Silva AC, Mak RH, Martelli DB, Silva LR, et al. Clinical characteristics and risk factors for death among hospitalised children and adolescents with COVID-19 in Brazil: an analysis of a nationwide database. Lancet Child Adolesc Heal 2021;5:559–68.

[19] Sousa BLA, Brentani A, Costa Ribeiro CC, Dolhnikoff M, Grisi SJFE, Ferrer APS, et al. Non-communicable diseases, sociodemographic vulnerability and the risk of mortality in hospitalised children and adolescents with COVID-19 in Brazil: a cross-sectional observational study. BMJ Open 2021;11:e050724.

[20] Chen JT, Krieger N. Revealing the unequal burden of COVID-19 by income, race/ethnicity, and household crowding: US county versus zip code analyses. J Public Heal Manag Pract 2021;27:S46–56.

[21] Dong E, Du H, Gardner L. An interactive web-based dashboard to track COVID-19 in real time. Lancet Infect Dis 2020;20:533–4.

[22] Peta Sebaran COVID-19. https://covid19.go.id/peta-sebaran (accessed 19 June 2022).

[23] Dyer O. Covid-19: Indonesia becomes Asia’s new pandemic epicentre as delta variant spreads. BMJ 2021;374:n1815.

[24] Surendra H, Salama N, Lestari K, Adrian V, Nurhasim A, Shankar AH, et al. Pandemic inequity in a megacity: a multilevel analysis of individual, community, and health care vulnerability risks for COVID-19 mortality in Jakarta, Indonesia. BMJ Glob Heal 2022;0:e008329.

[25] Surendra H, Elyazar IRF, Djaafara BA, et al. Clinical characteristics and mortality associated with COVID-19 in Jakarta, Indonesia : A hospital-based retrospective cohort study. Lancet Reg Heal - West Pacific 2021;9:100108.

[26] Mboi N, Murty Surbakti I, Trihandini I, et al. On the road to universal health care in Indonesia, 1990–2016: a systematic analysis for the Global Burden of Disease Study 2016. Lancet 2018;392:581–91.

[27] World Health Organization. Noncommunicable Disease (NCD) Country Profile, 2018. Geneva: 2018.

[28] Badan Pusat Statistik Indonesia. https://www.bps.go.id/dynamictable/2020/02/17/1771/indeks-pembangunan-manusia-menurut-kabupaten-kota-metode-baru-2010-2019.html 2019.

[29] Clark A, Jit M, Warren-Gash C, et al. Global, regional, and national estimates of the population at increased risk of severe COVID-19 due to underlying health conditions in 2020: a modelling study. Lancet Glob Heal 2020;8:e1003–17.

[30] Mahendradhata Y, Trisnantoro L, Listyadewi S, et al. Health Systems in Transition Vol. 7 No. 1 2017. The Republic of Indonesia Health System Review. vol. 7. 2017.

[31] Kementerian Kesehatan Republik Indonesia. Indeks Pembangunan Kesehatan Masyarakat.

[32] von Elm E, Altman DG, Egger M, et al. The Strengthening the Reporting of Observational Studies in Epidemiology (STROBE) statement: guidelines for reporting observational studies. J Clin Epidemiol 2008;61:344–9.

[33] Wirawan GBS, Januraga PP. Correlation of Demographics, Healthcare Availability, and COVID-19 Outcome: Indonesian Ecological Study. Front Public Heal 2021;9:1–8.

[34] Tchicaya A, Lorentz N, Leduc K, et al. COVID-19 mortality with regard to healthcare services availability, health risks, and socio-spatial factors at department level in France: A spatial cross-sectional analysis. PLoS One 2021;16:1–19.

[35] Sinto R, Utomo D, Suwarti, et al. Serum anti-Spike antibody titers before and after heterologous booster with mRNA-1273 SARS-CoV-2 vaccine following two doses of inactivated whole-virus CoronaVac vaccine. MedRxiv 2021.

[36] Pramana C, Indriana G, Setyopambudi K. Health Workers and Doctors Death During the Covid-19 Pandemic in Indonesia. Int J Med Rev Case Reports 2021;5:1.

[37] Harrison SL, Fazio-Eynullayeva E, Lane DA, et al. Comorbidities associated with mortality in 31,461 adults with COVID-19 in the United States: A federated electronic medical record analysis. PLoS Med 2020;17:1–11.

[38] Docherty AB, Harrison EM, Green CA, et al. Features of 20 133 UK patients in hospital with covid-19 using the ISARIC WHO Clinical Characterisation Protocol: Prospective observational cohort study. BMJ 2020;369:1–12.

[39] Hawkins RB, Charles EJ, Mehaffey JH. Socio-economic status and COVID-19–related cases and fatalities. Public Health 2020;189:129–34.

[40] Calderón-Larrañaga A, Vetrano DL, Rizzuto D, et al. High excess mortality in areas with young and socially vulnerable populations during the COVID-19 outbreak in Stockholm Region, Sweden. BMJ Glob Heal 2020;5.

[41] Victora PC, Castro PMC, Gurzenda S, et al. Estimating the early impact of vaccination against COVID-19 on deaths among elderly people in Brazil: analyses of routinely-collected data on vaccine coverage and mortality. EClinicalMedicine 2021;38:101036.

[42] Vaksinasi COVID-19 Nasional. Available: https://vaksin.kemkes.go.id/#/vaccines (accessed 19 June 2022).

