## Supplementary Table for "Heterogeneity and district-level factors associated with COVID-19 mortality during three epidemic waves in Indonesia: a nationwide ecological study"

**Supplementary data**

**Supplementary Table 1. COVID-19 incidence rate between March 2020 and February 2022 in Indonesia by sex and province**

| Province | Male | | | Female | | | Male-female Incidence rate ratio |
|---|---|---|---|---|---|---|---|
| | No. of cases | No. of population | Incidence rate | No. of cases | No. of Population | Incidence rate | |
| Aceh | 19,279 | 2,691,800 | 716.21 | 20,991 | 2,696,300 | 778.51 | 0.92 |
| Bali | 76,221 | 2,221,400 | 3431.22 | 75,737 | 2,193,100 | 3453.42 | 0.99 |
| Banten | 131,076 | 6,557,900 | 1998.75 | 133,673 | 12,895,300 | 1036.60 | 1.93 |
| Bengkulu | 12,046 | 1,015,200 | 1186.56 | 14,385 | 1,994,300 | 721.31 | 1.65 |
| Yogyakarta | 89,381 | 1,935,400 | 4618.22 | 100,757 | 3,919,200 | 2570.86 | 1.80 |
| Jakarta | 567,947 | 5,267,800 | 10781.48 | 588,849 | 10,576,400 | 5567.58 | 1.94 |
| Gorontalo | 6112 | 593,500 | 1029.82 | 6738 | 1,186,300 | 567.98 | 1.81 |
| Jambi | 15,330 | 1,831,300 | 837.11 | 16,917 | 3,604,200 | 469.37 | 1.78 |
| West Java | 480,932 | 25,111,200 | 1915.21 | 508,967 | 49,565,200 | 1026.86 | 1.87 |
| Central Java | 259,451 | 17,237,300 | 1505.17 | 294,624 | 34,738,200 | 848.13 | 1.77 |
| East Java | 252,742 | 19,722,200 | 1281.51 | 274,625 | 39,955,900 | 687.32 | 1.86 |
| West Kalimantan | 25,549 | 2,591,400 | 985.91 | 27,000 | 5,104,900 | 528.90 | 1.86 |
| South Kalimantan | 41,090 | 2,156,700 | 1905.23 | 40,388 | 4,268,600 | 946.17 | 2.01 |
| Central Kalimantan | 25,767 | 1,394,900 | 1847.23 | 26,179 | 2,686,300 | 974.54 | 1.90 |
| East Kalimantan | 104,211 | 1,902,900 | 5476.43 | 82,868 | 3,664,700 | 2261.25 | 2.42 |
| North Kalimantan | 20,546 | 374,100 | 5492.11 | 17,896 | 708,400 | 2526.25 | 2.17 |
| Bangka Belitung Islands | 27,093 | 756,900 | 3579.47 | 31,381 | 1,469,800 | 2135.05 | 1.68 |
| Riau Islands | 33,042 | 1,179,000 | 2802.55 | 30,594 | 2,309,500 | 1324.70 | 2.12 |
| Lampung | 28,001 | 4,364,300 | 641.59 | 36,645 | 8,534,800 | 429.36 | 1.49 |
| Maluku | 8,915 | 900,400 | 990.12 | 9,335 | 1,787,100 | 522.35 | 1.90 |
| North Maluku | 6,740 | 637,300 | 1057.59 | 6,519 | 1,252,300 | 520.56 | 2.03 |
| West Nusa Tenggara | 17,385 | 2,563,900 | 678.07 | 17,731 | 5,225,900 | 339.29 | 2.00 |
| East Nusa Tenggara | 34,252 | 2,731,600 | 1253.92 | 39,283 | 5,513,400 | 712.50 | 1.76 |
| Papua | 23,136 | 1,777,700 | 1301.46 | 20,842 | 3,393,100 | 614.25 | 2.12 |
| West Papua | 15,299 | 518,100 | 2952.91 | 13,751 | 986,000 | 1394.63 | 2.12 |
| Riau | 69,698 | 3,553,200 | 1961.56 | 71,452 | 6,951,200 | 1027.91 | 1.91 |
| West Sulawesi | 6,151 | 692,200 | 888.62 | 7,567 | 1,378,100 | 549.09 | 1.62 |

| | | | | | | | |
|---|---|---|---|---|---|---|---|
| South Sulawesi | 60,796 | 4,348,500 | 1398.09 | 73,206 | 8,888,800 | 823.58 | 1.70 |
| Central Sulawesi | 23,262 | 1,565,100 | 1486.30 | 29,149 | 3,081,700 | 945.87 | 1.57 |
| Southeast Sulawesi | 11,861 | 1,352,900 | 876.71 | 12,598 | 2,703,500 | 465.99 | 1.88 |
| North Sulawesi | 22,933 | 1,279,600 | 1792.20 | 25,371 | 2,512,900 | 1009.63 | 1.78 |
| West Sumatera | 42,875 | 2,760,600 | 1553.10 | 54,651 | 5,545,700 | 985.47 | 1.58 |
| South Sumatera | 36,933 | 4,358,000 | 847.48 | 39,159 | 8,600,800 | 455.29 | 1.86 |
| North Sumatera | 64,778 | 7,392,700 | 876.24 | 74,593 | 14,798,400 | 504.06 | 1.74 |

### Supplementary Table 2. Correlation matrix between each district-level variable assessed in this study

| | Mortality rate | Incidence rate | Elder population | Hypertension | Diabetes mellitus | Obesity | Pneumonia | Vaccine coverage | Doctor ratio | Nurse ratio | Midwife ratio | Hospital ratio | Expenditure | Life expectancy | Education |
|---|---|---|---|---|---|---|---|---|---|---|---|---|---|---|---|
| Mortality rate | | | | | | | | | | | | | | | |
| Incidence rate | 0.0000 | | | | | | | | | | | | | | |
| Elder population | 0.0000 | 0.0169 | | | | | | | | | | | | | |
| Hypertension | 0.0000 | 0.0000 | 0.0000 | | | | | | | | | | | | |
| Diabetes mellitus | 0.0000 | 0.0000 | 0.0000 | 0.0007 | | | | | | | | | | | |
| Obesity | 0.0000 | 0.0000 | 0.0307 | 0.8458 | 0.0000 | | | | | | | | | | |
| Pneumonia | 0.0316 | 0.0179 | 0.0001 | 0.8889 | 0.2477 | 0.2400 | | | | | | | | | |
| Vaccine coverage | 0.0000 | 0.0000 | 0.0000 | 0.0000 | 0.0000 | 0.0000 | 0.0000 | | | | | | | | |
| Doctor ratio | 0.0147 | 0.0000 | 0.0019 | 0.0007 | 0.0010 | 0.0010 | 0.5070 | 0.4155 | | | | | | | |
| Nurse ratio | 0.7569 | 0.0000 | 0.0000 | 0.0001 | 0.6374 | 0.0255 | 0.1832 | 0.0003 | 0.0000 | | | | | | |
| Midwife ratio | 0.0000 | 0.0000 | 0.4821 | 0.6588 | 0.2270 | 0.3261 | 0.1593 | 0.0000 | 0.0000 | 0.0005 | | | | | |
| Hospital ratio | 0.0000 | 0.0000 | 0.3757 | 0.0342 | 0.0000 | 0.0000 | 0.5500 | 0.0000 | 0.0000 | 0.0000 | 0.0000 | | | | |
| Expenditure | 0.0000 | 0.0000 | 0.0007 | 0.0010 | 0.0000 | 0.0000 | 0.0027 | 0.0000 | 0.0045 | 0.8661 | 0.1863 | 0.0000 | | | |
| Life expectancy | 0.0000 | 0.0000 | 0.0000 | 0.0000 | 0.0000 | 0.0000 | 0.2332 | 0.0000 | 0.2701 | 0.0003 | 0.0032 | 0.0000 | 0.0000 | | |
| Education | 0.0000 | 0.0000 | 0.0384 | 0.0020 | 0.0000 | 0.0000 | 0.7313 | 0.0000 | 0.0000 | 0.0000 | 0.0002 | 0.0000 | 0.0000 | 0.0000 | |
